# Fear of Infection and Sufficient Vaccine Reservation Information Might Drive Rapid Coronavirus Disease 2019 Vaccination in Japan: Evidence from Twitter Analysis

**DOI:** 10.1101/2022.05.15.22275071

**Authors:** Qian Niu, Junyu Liu, Masaya Kato, Tomoki Aoyama, Momoko Nagai-Tanima

## Abstract

**Background:** The global public health and socioeconomic impacts of coronavirus disease 2019 (COVID-19) have been substantial, making herd immunity by COVID-19 vaccination an important factor for protecting people and retrieving the economy. Among all the countries, Japan became one of the countries with the highest COVID-19 vaccination rate in several months, although the vaccine confidence in Japan is the lowest worldwide.

**Objective:** We attempted to find the reasons for the rapid coronavirus disease 2019 (COVID-19) vaccination in Japan under the lowest vaccine confidence in the world by Twitter analysis.

**Materials and methods:** We downloaded COVID-19 related Japanese tweets from a large-scale public COVID-19 Twitter chatter dataset within the timeline of February 1, 2021 and September 30, 2021. The daily number of vaccination cases was collected from the official website of the Prime Minister’s Office of Japan. After preprocessing, we applied unigram and bigram token analysis, then calculated the cross correlation and Pearson correlation coefficient (*r*) between the term frequency and daily vaccination cases. Then we identified vaccine sentiments and emotions of tweets and used the topic modeling to look deeper into the dominant emotions.

**Results:** We selected 190,697 vaccine-related tweets after filtering. By n-gram token analysis, we discovered the top unigrams and bigrams over the whole period. In all the combinations of the top six unigrams, tweets with both keywords “reserve” and “venue” showed the largest *r* = 0.912 (P < 0.001) with the daily vaccination cases. In sentiment analysis, negative sentiment overwhelmed positive sentiment, and fear was the dominant emotion across the period. For the latent Dirichlet allocation model on tweets with fear emotion, the two topics were identified as “infect” and “vaccine confidence”. The expectation of the number of tweets generated from topic “infect” was larger than “vaccine confidence.”

**Conclusion:** Our work indicated that awareness of the danger of COVID-19 might increase the willingness to get vaccinated; With sufficient vaccine supply, effective vaccine reservation information delivery may be an important factor for people to get vaccinated; We didn’t find evidence for increased vaccine confidence in Japan during the period in our research. We recommend policymakers to share fair and prompt information about the infectious diseases and vaccination, and make efforts on smoother delivery of vaccine-reservation information.

## 1. Introduction

Coronavirus disease 2019 (COVID-19) has spread worldwide since its first case in December 2019 and has become a public health emergency of international concern [1]. Until September 30, 2021, Japan experienced five waves of the COVID-19 pandemic [2]. The surge of COVID-19 in Japan occurred during the Tokyo Olympics, bringing the cumulative number of COVID-19 cases to 1,556,998 when the Games finished. However, with the lifting of the fourth national state of emergency on September 30, 2021, the nationwide pandemic was effectively contained, and the number of new confirmed cases abruptly decreased. The high vaccination rate in Japan was considered an important factor in stopping the fifth wave [3].

A high vaccination rate is thought to be promoted by high vaccine confidence [4]. Vaccine confidence is the belief that vaccines work, are safe, and are part of a trustworthy medical system [5]. A global survey that did not include Japan showed that the potential acceptance of a COVID-19 vaccine largely varied among countries [6]. Japan ranks among the countries with the lowest vaccine confidence worldwide according to a survey in 2020 [7]. Another survey conducted before the large-scale vaccination in Japan indicated that Japan ranked last in confidence in COVID-19 vaccines among 15 countries [8]. Gordon and Reich explained the historical reasons for low vaccine confidence in Japan [9]. Kunitoki et al. proposed that barriers to vaccine access and use are mainly from effective public communication and called for rebuilding vaccine confidence in Japan [10].

However, Japan’s speed of vaccination has been impressive since the large-scale vaccination (May 24, 2021). Japan’s first dose vaccination rate was approximately 6.8% by June 1, 2021, and over 70% of the population accepted at least one dose until September 30, 2021 [11]. Notably, vaccination was not mandatory and was administered only with the recipient’s consent. A survey of multiple countries reported the coexistence of a high level of uncertainty about the safety of COVID-19 vaccines and a high willingness to get vaccinated [12], which indicates that Japan may not be a special case. The reason for the contradiction between the rapid growth of the COVID-19 vaccination rate and low vaccine confidence in Japan is worth studying and maybe instructive for propelling worldwide vaccination towards infectious diseases.

Twitter is a widespread social media platform that has attracted the increasing attention of public health researchers because of its advantages of large amounts, real-time availability, and ease of public searching and accessing [13]. With a large amount of real-time COVID-19-related posts, Twitter has been widely used for public opinion mining towards COVID-19 during the pandemic, providing policymakers with substantiated evidence [12,14,15]. Lyu et al. reported the trend of topics and sentiments of English tweets for approximately 11 months since the World Health Organization declared the COVID-19 pandemic [14]. Yousefinaghani et al. reported the dominance of positive sentiments and more vaccine objection and hesitancy than vaccine interest [12]. Huangfu et al. reported the results of topic modeling and sentiment analysis of tweets between December 8, 2020 and April 8, 2021 [15]. Eibensteiner et al. reported willingness to vaccinate despite the safety concerns of vaccines, according to a survey on Twitter Poll [16]. Besides, Twitter is the most popular social media platform in Japan [17], owning 58.2 million users as of October 2021[18], making Twitter analysis more powerful for COVID-19 research in Japan. A Twitter analysis by Niu et al. reported that the Japanese public’s negative sentiment overwhelmed the positive sentiment towards the COVID-19 vaccine before and at the beginning of the large-scale vaccination campaign [19].

This retrospective study aimed to identify the reasons for the rapid COVID-19 vaccination in Japan by analyzing tweets. We hypothesized that the increase in vaccination rates might be due to subjective factors, including S1) increased public confidence in vaccines and S2) fear of infection, and objective factors, including O1) adequate vaccine supply and O2) effective delivery reservation-related vaccine information. To test these hypotheses, we collected vaccine-related tweets posted between February 1, 2021, and September 30, 2021. Then, we preprocessed the collected tweets and conducted a unigram token analysis, sentiment analysis, and topic modeling.

## 2. Method

In previous works of large-scale Twitter analyses, after preprocessing, there are mainly four types of NLP methods: n-gram token analysis [12,15,20,21], sentiment analysis [12,14,15,20– 25], topic modelling [12,14,15,20,22–25], and geographical analysis [22,24]. The geographical analysis is less important in our work because the range of our research is a whole country instead of subareas. In this work, we followed the previous works in applying the n-gram token analysis, sentiment analysis, and topic modeling.

### 2.1 Data collection and preprocessing

The data used in this study were obtained from a large-scale public COVID-19 Twitter chatter dataset [26] updated by the Georgia State University’s Panacea Lab. The dataset provided the identifications (IDs), posting time, and the languages of all the tweets were provided in the dataset. We downloaded COVID-related Japanese tweets between February 1, 2021, the month the first person was vaccinated, and September 30, 2021, when the first-dose vaccination rate exceeded 70%. In addition, data on the number of vaccination cases were collected from the official website of the Prime Minister’s Office of Japan (PMOJ) [27].

The downloaded tweets were then cleaned and processed. Retweets were filtered using the Python package tweepy. Tweets that included no keywords related to vaccines were deleted. It worth notice that the three vaccine brands (Pfizer, Moderna, and AstraZeneca) that were approved by the Japanese government were included in the keywords. Other vaccine brands were excluded because we tried to focus more on the brands adopted in the vaccination process. Frequent misspellings (e.g. Modelna) was also included in the keywords. Weblinks, special characters, emojis, and “amp” (ampersands) were removed, and all full-width English characters were transferred to half-width and lowercase.

For convenience, all Japanese words in our results were directly presented in English translations. In order to minimize the influence of difference between languages, all the translations in our results were done as the last step by directly replacing the Japanese words in the graphs with corresponding English words, and therefore won’t influence the statistical results.

### 2.2 Unigram and bigram token analysis

Tokenization is necessary before many other natural language processing (NLP) tasks, especially for many non-Latin languages, such as Japanese. We removed the pre-defined English, and Japanese stop words in the Python packages NLTK [28] and SpaCy [29], and tokenized all collected vaccine-related tweets using the Python package SpaCy into unigrams or bigrams for statistical analysis, as in the work of Kwok et al. [26]. We sorted the unigram tokens or bigram tokens in descending order of term frequency over the entire period. Similar to Liu et al. [24], we used the pruned exact linear time (PELT) algorithm [30] to find the first change point of the term frequency. Unigrams before the first change point were regarded as top unigrams, and the term frequencies of the unigrams after the change point were significantly lower than the top unigrams. Similar processes were done for bigrams. To eliminate the difference in the number of days between months, the monthly term frequency was defined by dividing the total term frequency by the number of days each month for each top unigram or bigram.

Correlation coefficients were widely used in social media analysis. In Google Trends analysis, correlations were calculated between reported cases of infectious disease and the trends of search for relevant keywords. In Twitter analyses, correlations between the daily infectious or death cases, and the number of related tweets or sentiment scores, were also investigated [24,25]. In this work, correlation analyses were adopted to find out the factors from the top unigrams that are most related to COVID-19 vaccination campaign. we first calculated the cross correlations between the number of tweets containing the top unigrams or bigrams and the vaccination cases, then observed the time lags when maximum cross correlation appeared for each unigram and bigram. Pearson correlation coefficients (*r*) between top unigrams or bigrams and the vaccination cases were also calculated.

### 2.3 Sentiment analysis

After n-gram analysis, sentiment analyses were often used to explore the real-time public attitudes in social media analysis related to COVID-19 vaccination, which may reflect the acceptance of COVID-19 vaccines and related policies [12,14,20,22,24]. Trend of negative sentiments may provide potential evidence for vaccine hesitancy [23]. In this work, sentiment analysis was applied to all vaccine-related tweets. Cloud services were used in this study because there were no reliable public models for sentiment analysis in the Japanese language. We selected the Amazon Web Service (AWS) for consistency with previous work [28]. The tweets were divided into positive, negative, neutral, or a mixture of positive and negative tweets using the AWS. Fine-grained emotions were also explored using the Japanese version of the NRC Emotion Lexicon [31].The NRC Emotion Lexicon is a dictionary of words and their associated scores for eight emotions: anticipation, trust, joy, surprise, anger, disgust, fear, and sadness. The positive and negative tweets were tokenized, and the degree of valence (DOV) for the eight emotions was calculated by adding up the scores for the unigrams that appeared in the NRC Emotion Lexicon. Finally, we calculated the daily average DOV by dividing the number of positive and negative tweets on that day to show the trend of each emotion.

### 2.4 Topic modeling

Topic modeling were applied to identify fine-grained information from tweets of different sentiments [12,15,24]. Based on the sentiment analysis results, we summarized the topics to look deeper into the dominant emotion in the tweets. Latent Dirichlet allocation (LDA) is often used in tweets topic modeling studies [14,15,20,22,23]. In this study, LDA regards tweets as being generated from different topics, and each topic generates tweets with a Dirichlet distribution. A Python package scikit-learn was used to determine the best number of topics. Log likelihood was adopted as the metric for selection, and five-fold cross correlation was applied to avoid overfitting. According to figure 1 in the Appendix, we chose two as the number of topics for LDA modelling, which showed highest log likelihood score. We used scikit-learn for the LDA topic modeling and displayed the top ten keywords and their weights related to each topic. The weights were the pseudo-counts of the keywords in a topic. The themes of topics were summarized from the top ten keywords by three volunteers. The volunteers were first asked to work out the themes of the topics independently, and then they had a meeting to finally reach an agreement on the themes.

We then checked the trends of tweets related to different topics. Defining the i-th tweet in all collected tweets as *d*_*i*_ and the j-th topic of the LDA model as *t*_*j*_, the probability of a tweet *d*_*i*_ coming from *t*_*j*_ was calculated using the fitted LDA model as *p*_*ij*_. For tweets posted each day, the expectation of the number of tweets generated from topic j was calculated by summing *p*_*ij*_ on that day. The ratio between the expected number of tweets generated from each topic was also plotted to show the trend of public attention under dominant emotion.

## 3. Results

### 3.1 Data summary

We downloaded 979,636 Japanese tweets posted between February 1, 2021, and September 30, 2021, according to the ID and region information in the dataset. After filtering, 190,697 vaccine-related tweets were selected. As a result, the total number of vaccine-related tweets increased from 14,758 tweets in February to 34,692 in August and then decreased to 27,824 in September.

### 3.2 Unigram and bigram token analysis

The change point of unigram term frequencies detected by the PELT algorithm was six, and the top six unigrams were Japanese words for “infection,” “Japan,” “reserve,” “Pfizer,” “venue,” and “mutation.” The unigram “side effects”, related to the safety of vaccines, ranked eighth overall. The unigrams “infect,” “reserve,” and “venue” gradually ranked in the top-3 from February to September, as shown in Fig. 1.

**Fig. 1.**
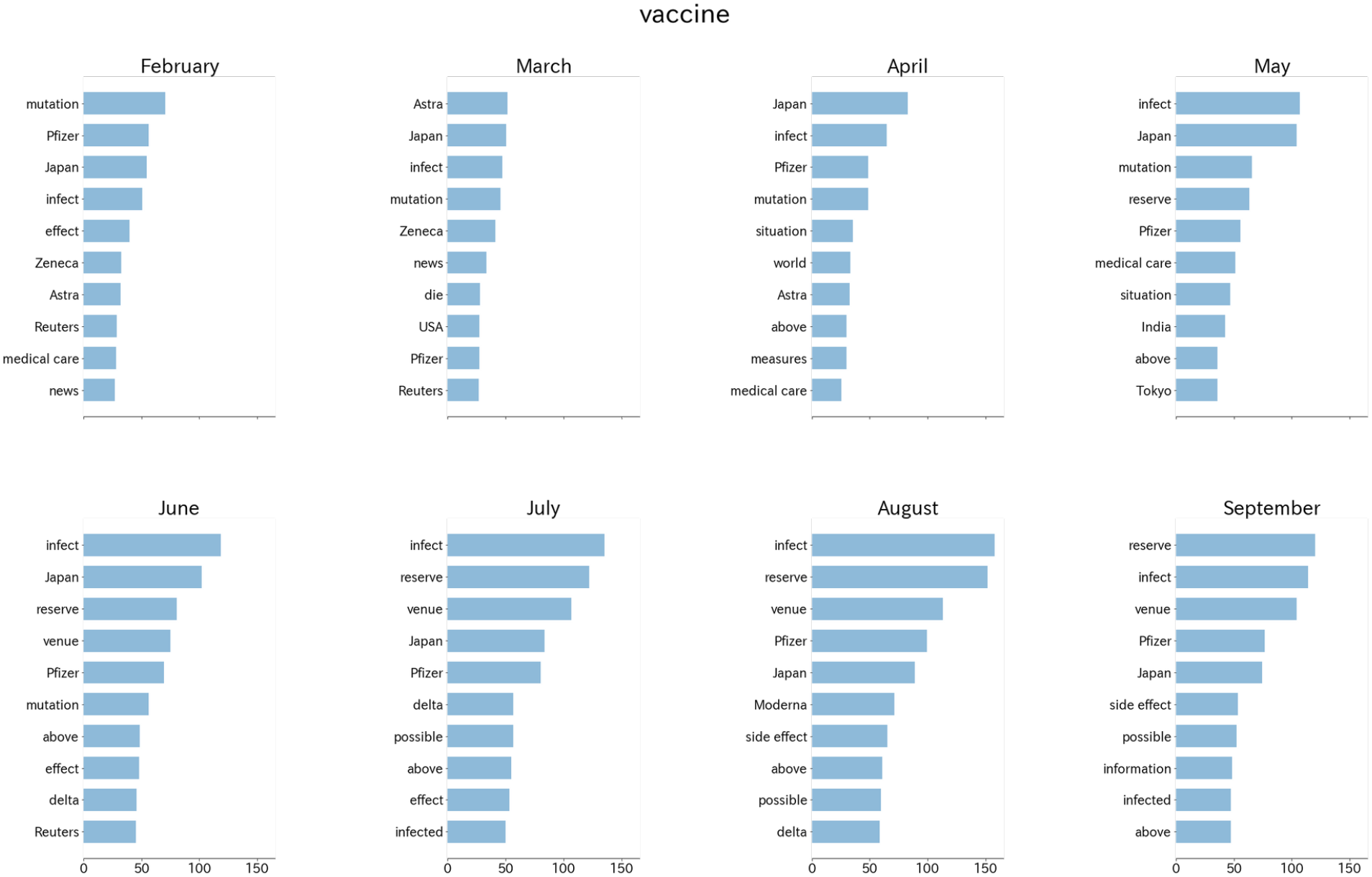
Translation of top ten unigrams of each month. The lengths of the bars represent the monthly term frequencies in tweets of each month.

The change point of bigram term frequencies detected by the PELT algorithm was five, and the top five were Japanese bigrams for “Astra + Zeneca,” “reserve + possible,” “article + Reuters,” “venue + reserve,” and “medical-care + workers.” The bigrams “reserve + possible” and “venue + reserve” ranked in the top-2 from June to September, and the ranking of “Astra + Zeneca” decreased since May, as shown in Fig. 2.

**Fig. 2.**
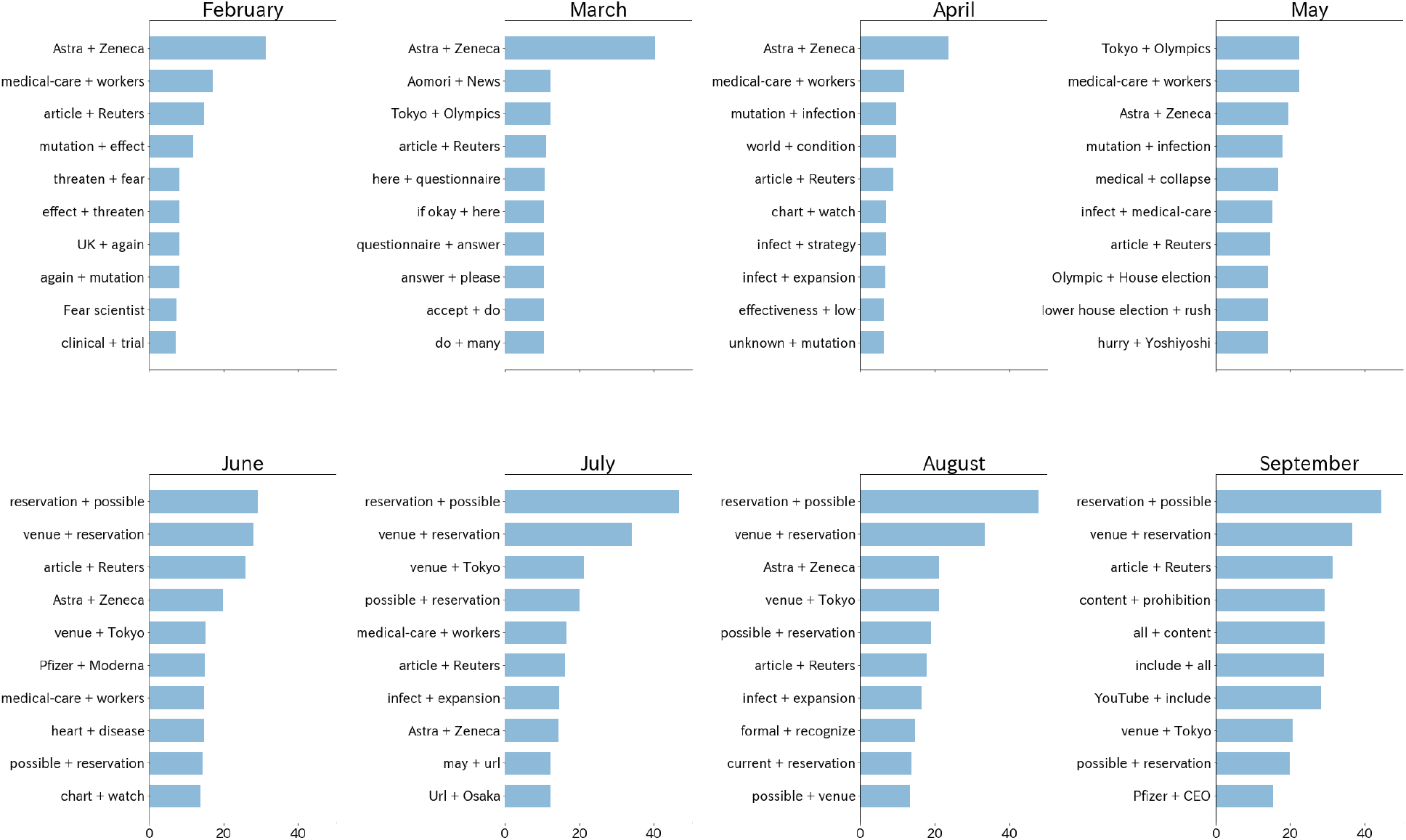
Translation of top ten bigrams of each month. The lengths of the bars represent the monthly term frequencies in tweets of each month.

As for correlation analysis of unigrams, the time lags for “reserve” and “venue” were 0, and the vaccination cases led the number of tweets containing “infection” for 5 days. After calculating *r* between the daily number of tweets containing each top unigram and vaccination cases, significant *r* (*P* < 0.001) was found for all unigrams except “mutation.” The largest *r* for the daily vaccination cases was from unigrams “infection” (*r* = 0.746), “reserve” (*r* = 0.829), and “venue” (*r* = 0.908). We then checked the daily number of tweets containing all the combinations of the three unigrams showing a strong correlation and found the highest *r* (*r* = 0.912, *P* < 0.001) for tweets containing both “reserve” and “venue.” By randomly selecting 5 days and checking the source of all the tweets on those days, we found the percentage of tweets containing both “reserve” and “venue” posted by official accounts or mainstream media was 96.0%–100% (95% confidence interval [CI]). The trend of tweets containing both unigrams “reserve” and “venue” compared with the daily vaccination cases is shown in Fig. 3.

**Fig. 3.**
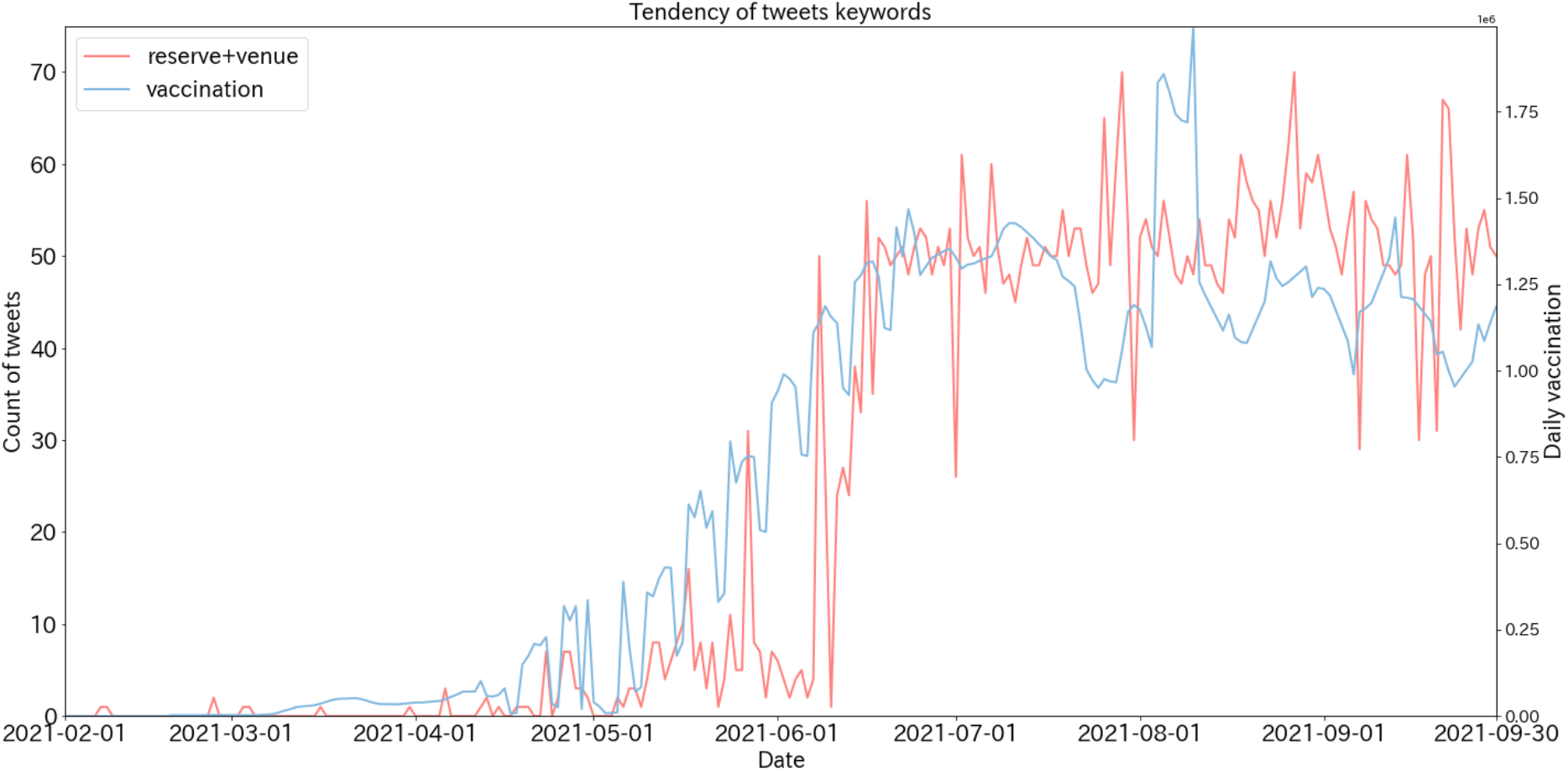
Trend of tweets containing both unigrams “reserve” and “venue,” and the curve for daily first dose vaccination cases

As for bigrams, the bigram “venue + reserve” overlapped with the unigram analysis and was excluded from this part. The time lags for bigrams “reserve + possible” and “article + Reuters” were 0, and vaccination cases led “Astra + Zeneca” and “medical-care + workers” for 116 days and 63 days respectively. The bigrams “reserve + possible” and “article + Reuters” got the highest cross correlations than the rest. The bigrams “Astra + Zeneca” (*r*=-0.331), “reserve + possible” (*r*=0.908), and “article + Reuters” (*r*=0.229) showed significant *r* (*P*<0.001) except “medical-care + workers” (*r*=-0.055). By three volunteers’ manually checking, we found 95.4% of the tweets contains the bigrams “reserve + possible” were the same as those of the combination of unigrams “venue” and “reserve”.

### 3.3 Sentiment analysis

For all tweets, 4,453 (2.3%) were positive, 19,340 (10.1%) were negative, 164,687 (86.4%) were neutral, and 2,217 (1.2%) were mixed positive and negative sentiments. A comparison between the daily numbers of tweets marked as positive and negative is shown in Fig. 4. Negative sentiments overwhelmed positive sentiments for all days.

**Fig. 4.**
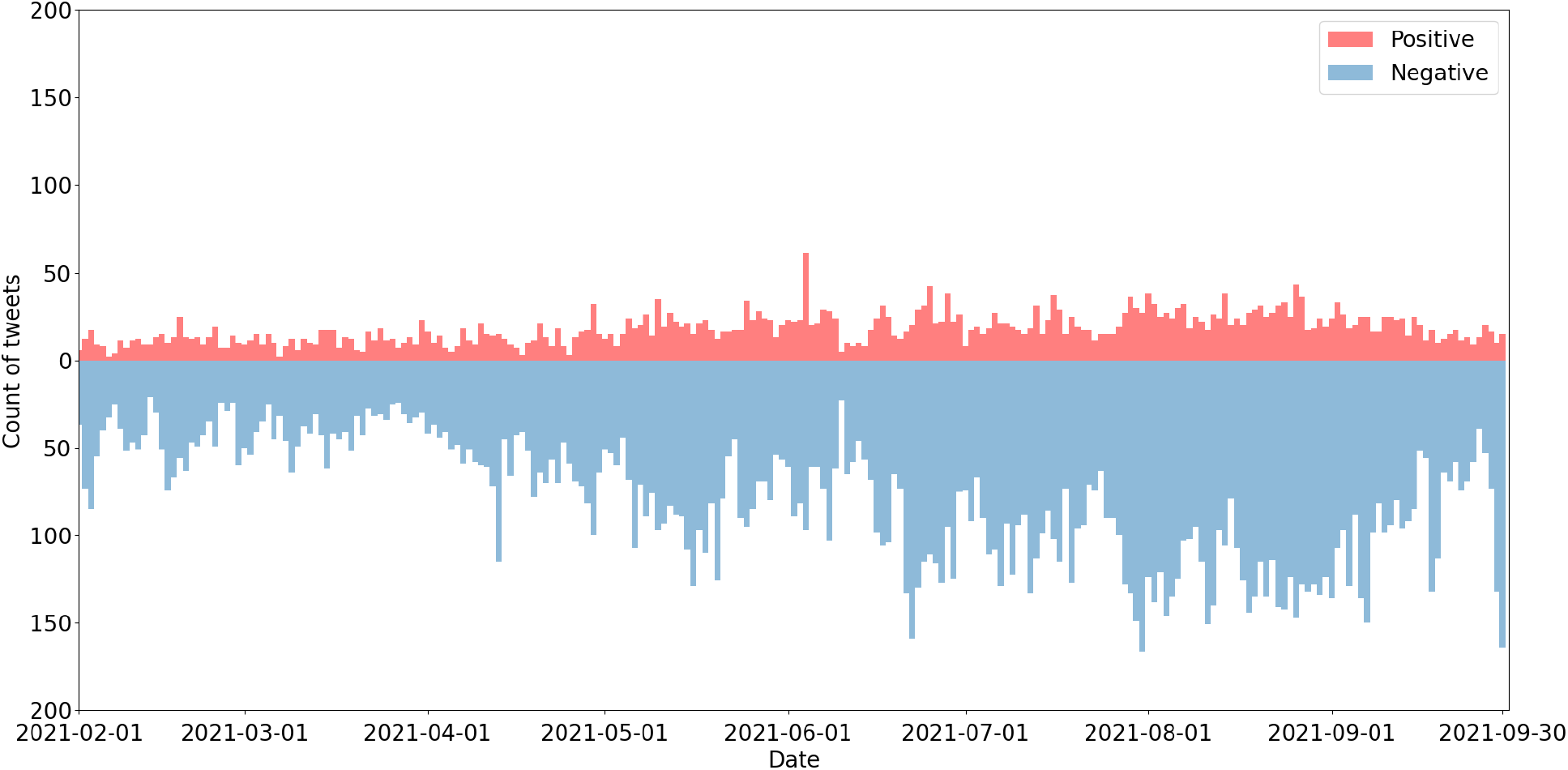
Comparison between the daily number of tweets marked positive (orange) and negative (green)

The DOVs for the eight emotions are shown in Fig. 5. The daily average DOV of anger (0.404), disgust (0.268), fear (0.659), sadness (0.486), overwhelmed anticipation (0.163), trust (0.173), joy (0.118), and surprise (0.081). Fear was the dominant emotion during this period. Here, we defined the peaks of emotion as larger than three times the daily average DOV for that emotion. Trust peaked (1.114) on February 18, 2021. From May 13, 2021 to May 18, 2021, there were several peaks of fear.

**Fig. 5.**
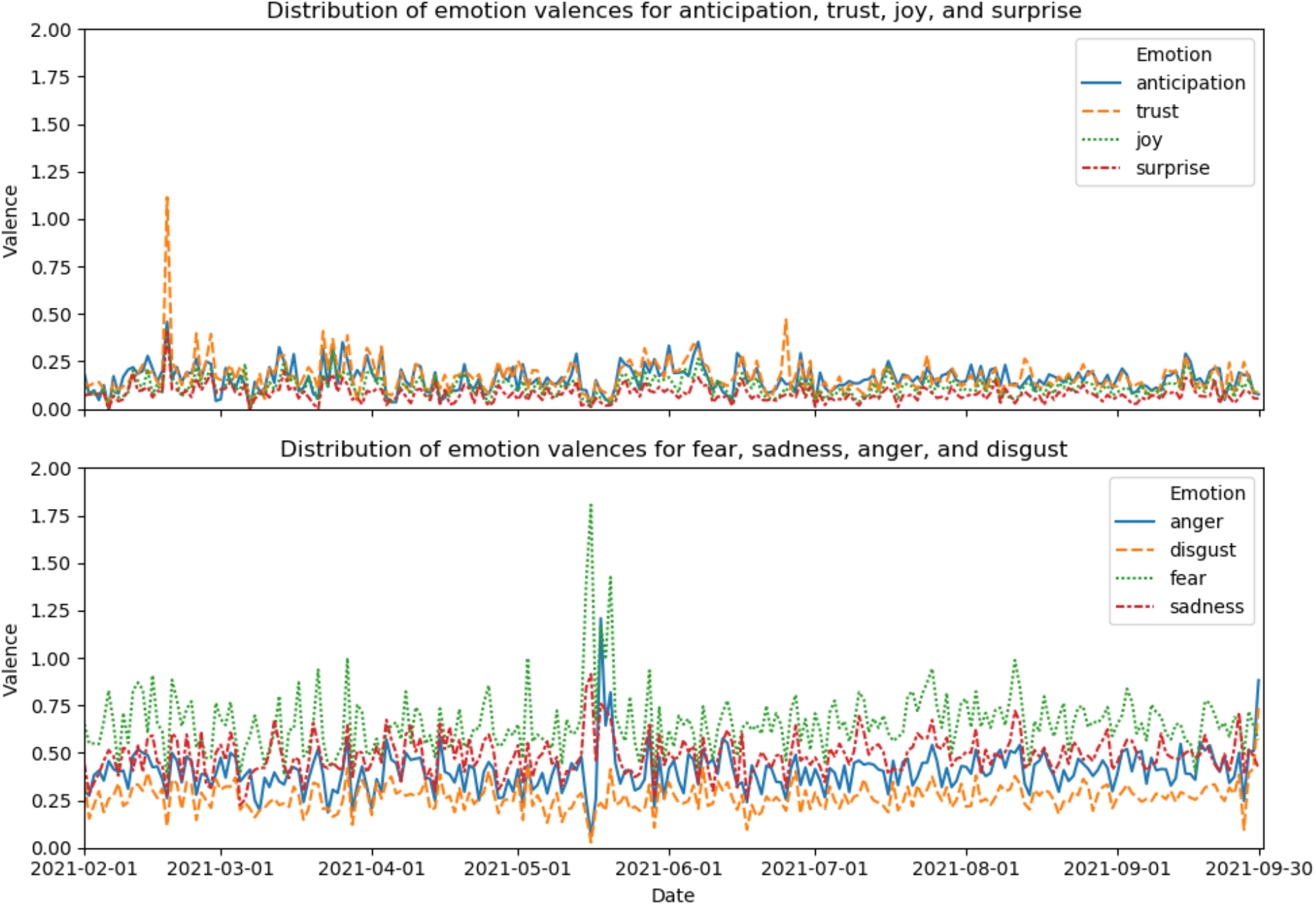
Daily average DOV of eight emotions in the vaccine-related tweets

### 3.4 Topic modeling

The top ten keywords for each LDA topic are shown in Fig. 6. The theme of topic-1 is “infect,” and topic-2 is “vaccine confidence.” It is also noticeable that the weight of “infect” (14,895) in topic-1 was over three times the weight of the second keyword “Japan” (4,359), but the weight of “Pfizer” (4,348) in topic-2 was only 15.5% larger than the second keyword “die” (3,763).

**Fig. 6.**
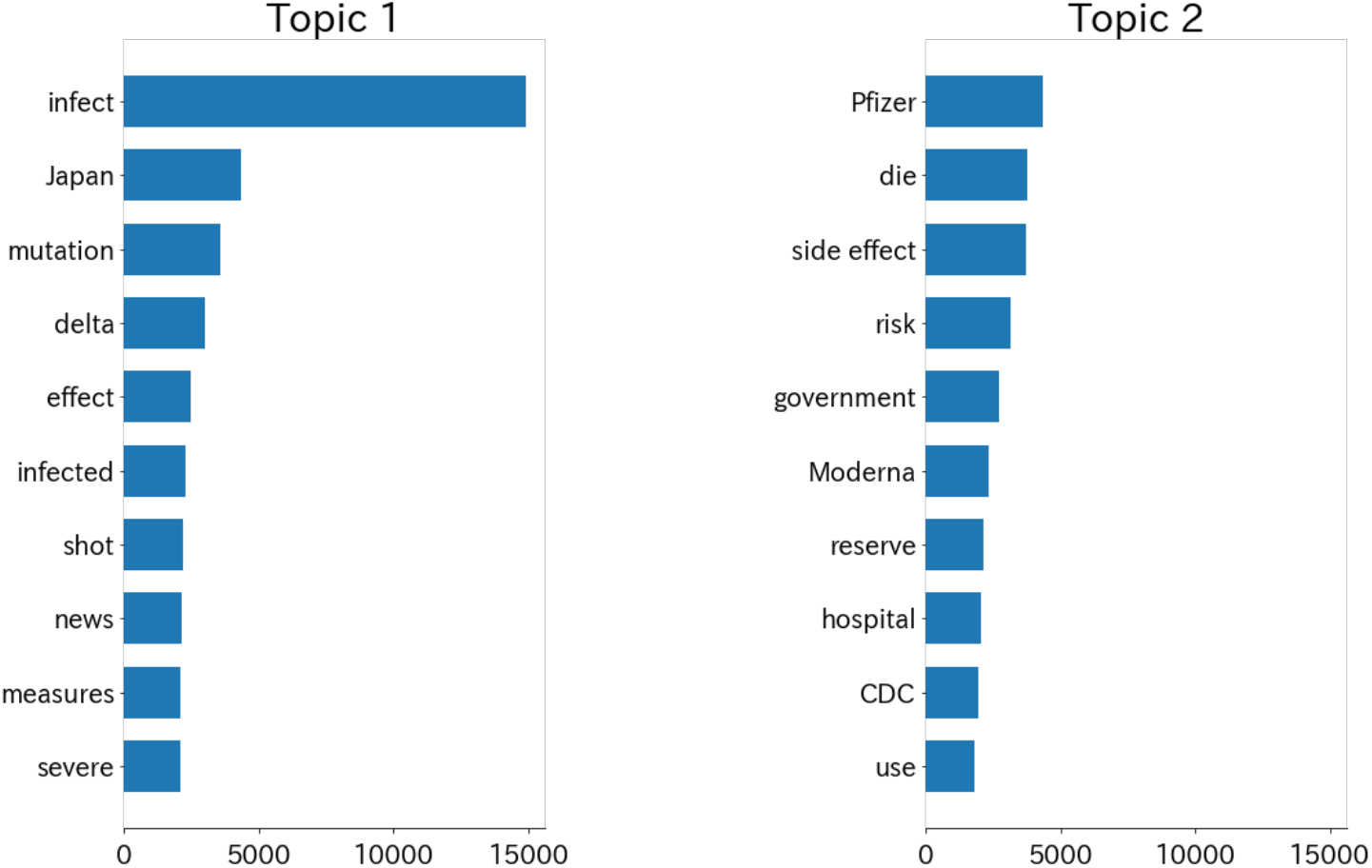
Top ten keywords of two topics by LDA modeling. The bars represent the weights, which can be regarded as the pseudo counts of the keywords in each topic.

The ratio between the expectation of the number of “infect”-related tweets and “vaccine confidence” vaccine confidence-related tweets is shown in Fig. 7. The total expectation of the number of tweets generated from topic-1 (“infect,” n=30,288) is larger than topic-2 (“vaccine confidence,” n=27,572), and the mean ratio between the expectation of the daily number of tweets generated from topics-1 and -2 is significantly larger than 1 (*P* < 0.01). On 68.2% of days, the expectation of the number of tweets generated from “infect” was larger than “vaccine confidence.”

**Fig. 7.**
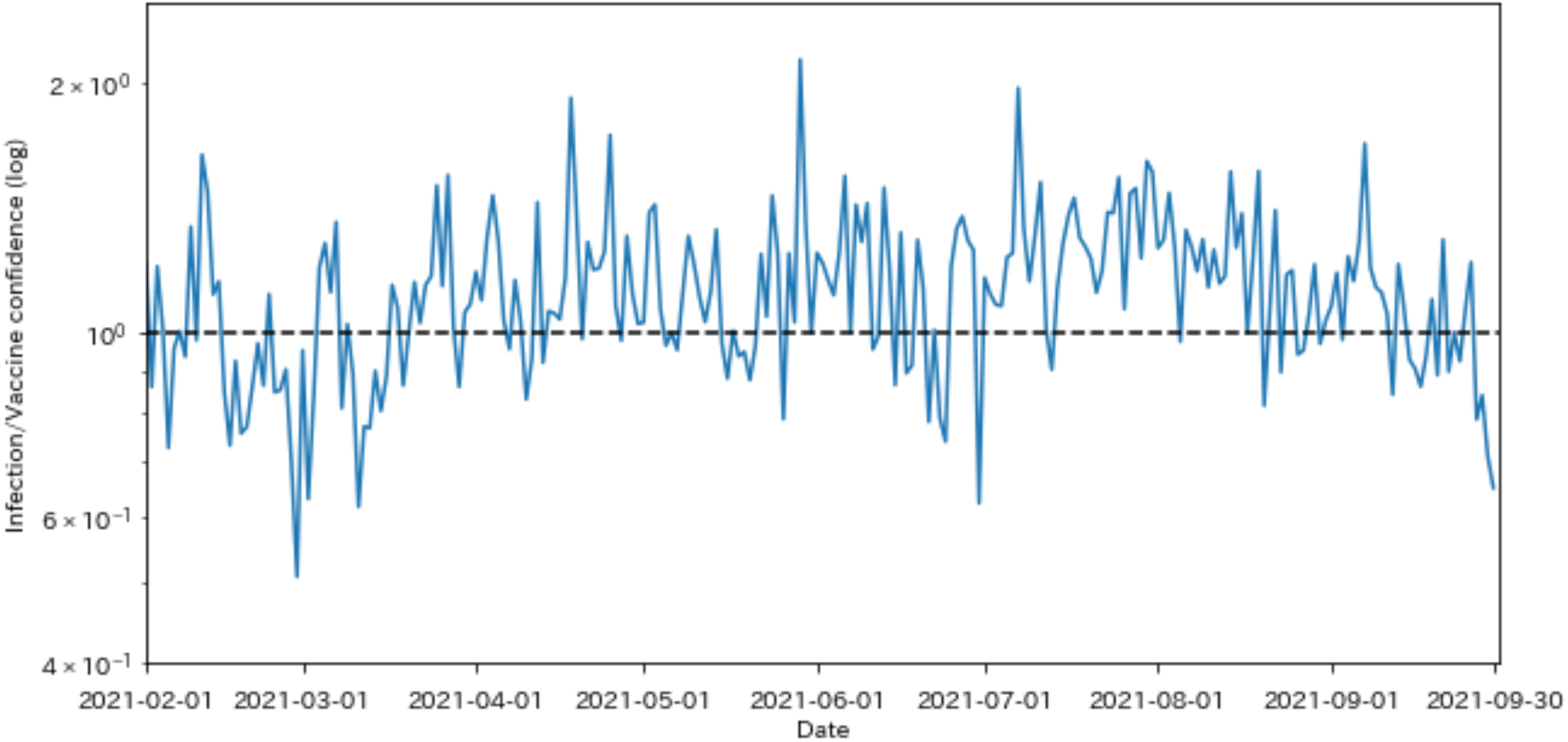
Ratio between the expectation of the number of “infection”-related tweets and “vaccine confidence”-related tweets

## 4. Discussion

A high vaccination rate is thought to be promoted by high vaccine confidence [16,36–39], but Japan achieved a high vaccination rate in several months, with the lowest vaccine confidence in the world. This retrospective study aimed to determine the reasons for the fast vaccination process in Japan, which may be instructive for propelling worldwide vaccination towards infectious diseases. Based on previous studies [16,37–40], we hypothesized that subjective factors, including S1) increased vaccine confidence and S2) fear of infection, and objective factors, including O1) adequate vaccine supply and O2) effective delivery of reservation-related vaccine information. Our results indicated that hypotheses S2 and O2 might have driven the public to be vaccinated. However, we did not find any evidence for hypothesis S2. Evidence for hypothesis O1 can be found in the history of vaccine supply on the official website of the Prime Minister’s Office of Japan (Prime Minister of Japan and his Cabinet) and is not discussed in this paper.

Several results support hypothesis S2. In Fig. 1 of the unigram token analysis, the keyword “infect” ranked the top three, except in February, and ranked first from May to August, during Japan’s fourth and fifth wave infections. The keywords “venue” and “reserve” also ranked up from May. No keywords related to increased vaccine confidence were found. Fig. 4 in sentiment analysis showed that negative sentiment overwhelmed positive, consistent with the results of Chen et al. that Japan showed dissatisfaction compared with neighboring countries [38]. Combined with our result that “infect” was the top-1 keyword and “side effect” ranked eighth in the unigram token analysis, our results support that the Japanese public was more concerned about infection than the side effects of COVID-19 vaccines.

More evidence for hypothesis S2 was obtained from the topic modeling results. From the keywords of topic-1 (“infect”), we can see that the public was concerned about the infection and death rate. The mutated virus and empowered cases also led to fear. Willis et al. found that less fear of infection may lead to a lower willingness to be vaccinated [40], which is complementary to our results. From the keywords of topic-2 (“vaccine confidence”), we can see that the side effects of the vaccines were the most concerning, but the following keywords were about the effectiveness of vaccines on the mutated virus, reservation of vaccines, and medical care conditions. Previous surveys in different countries have indicated that fear of vaccine safety is the key factor for low vaccine acceptance [41,42]. Also, the “side effect” weight in topic-2 was much less than “infect” in topic-1. The top keywords in the two topics indicated that people were more concerned about COVID-19 than the side effects of vaccines. Bendau et al. reported a significantly positive correlation between fears of infection and vaccine acceptance and a significant negative correlation between fear of vaccine safety and vaccine acceptance [43].Therefore, it is important to distinguish the mainstream fear emotion to determine the reason for the high vaccination rate. Fig. 7 provided details about the ratio between the expected number of tweets related to “infect” and “vaccine confidence.” In most cases, the ratio was larger than 1, indicating that the public was more concerned about infection than the safety and effectiveness of vaccines. Higher ratios were observed in April and from July to the end of September, periods of Japan’s fourth and fifth waves of infection. There was also a relatively long period of less than one ratio from mid-February to mid-March, which was the period when the vaccines were less effective against the mutated virus (February 10), severe side effects of the AstraZeneca vaccine (March 12), and several side effects in Japan (February 21, March 7, March 10). However, the ratio soon increased because of the fourth wave infection. This example also proved that fear of infection overcame the vaccine safety concern.

We also provide evidence of a strong relationship between vaccination and hypothesis O2. Bigram analysis in Fig 2 showed that “reservation + possible” ranked first since June, shortly after the large-scale vaccination started, which might reflect the strong concerns about the vaccine reservation by the public. Unigram token analysis in Fig. 3 showed that tweets including the keywords “reserve” and “venue” were significantly highly correlated (*r* > 0.9, *P* < 0.01) with the daily number of vaccination cases in Japan, and most of them were from government official accounts. The bigram “reservation + possible” also showed high correlation (*r* > 0.9, *P* < 0.01) with the daily vaccination cases. Because reservation information should always lead to the actual vaccination, this result indicated that in addition to sufficient vaccine supply, reservation information delivery might also be important in large-scale vaccination. Also, the time lag for the maximum cross correlation was 0, which may indicate the efficiency of the reservation information posted on Twitter. Our results were consistent with Fu’s work that inflexible information systems for vaccine reservation can impair immunization services in the community [44].

We did not find any evidence for hypothesis S1. Macaraan reported a shift from hesitancy to confidence towards the COVID-19 vaccination program among Filipinos [39]. Okubo [45] reported a shift from hesitancy to confidence, but they also admitted that the shift might come from the differences in metrics of the survey with previous work [46]. Following these works, we looked for a similar shift in sentiment or emotions from negative to positive, but negative sentiments overwhelm positive sentiments in Fig. 4, and fear dominates all the emotions in Fig. 5. The positive emotions, “anticipation,” “trust,” and “joy” did not increase during the entire period. These two results made it difficult to conclude increased vaccine confidence.

Our results were partially related to the 5 C model (confidence, competence, convenience, calculation, and collective responsibility) measuring vaccine hesitancy [39,47]. Confidence and complacency are two subjective measures that are directly related to individuals. In our work, the LDA theme “vaccine confidence” belonged to “confidence,” and “fear of infection” belonged to “complacency.” In Japan, fear of infection may drive a high vaccination rate. The delivery of reservation information may be an extension to “convenience,” which was previously defined as “physical availability, affordability and willingness-to-pay, geographical accessibility, ability to understand (language and health literacy), and appeal of immunization service affect uptake [48].” Our work indicates that information about vaccination reservations should also be considered for the convenience of vaccination.

We admit that our research might have some potential limitations: (1) the imbalance of the demographics of Twitter users in Japan [49] may cause bias in the results; (2) status of the user on a certain day (at home or not, other events on that day, etc.) may also bias the dataset [50]; (3) due to the lack of a reliable public model for sentiment analysis in the Japanese language, the cloud service AWS was used for sentiment analysis; (4) filtering keywords may include irrelevant or missing related tweets; (5) anti-vaccine tweets, especially rumors, were not distinguished or analyzed separately in this study. However, feature works can be combined with classical surveys to train the sentiment analysis model and model to distinguish rumors from tweets to overcome these limitations.

This retrospective study aimed to determine the reasons for the fast vaccination process in Japan, which might be instructive for propelling worldwide vaccination towards infectious diseases. In conclusion, our work indicated that awareness of the danger of COVID-19 increased the willingness to be vaccinated; with a sufficient supply of vaccines, effective reservation information delivery might provide more opportunities for people to be vaccinated. Models measuring vaccine hesitancy might also need to add efficiency in delivering reservation information as a metric. Based on our findings, we recommend public health policymakers and the government to share fair and prompt information about the infectious diseases and vaccination. Also, efforts on tied cooperation among multi-level relevant organizations and new media operation may lead to achieve smoother delivery of vaccine-reservation information.

## Data Availability

All data produced in the present study are available upon reasonable request to the authors

## Authors’ contribution

QN and JL performed analyses and drafted the manuscript. All authors conceived of the study, interpreted the results, and revised the manuscript. All authors have read and approved the final manuscript.

## Ethical Approval

This study used publicly available and accessible tweets collected by Georgia State University’s Panacea Lab allowing free download. We assert that our analysis is compliant with Twitter’s usage policy in aggregate form without identifying specific individuals who made the Twitter posts. Also, the numbers of infections, deaths, and vaccinated cases downloaded from the PMOJ are open government data. Therefore, the activities described do not meet the requirements of human subject research and did not require review by an institutional review board.

## Abbreviations

COVID-19: Coronavirus disease 2019
IDs: identifications
PMOJ: Prime Minister’s Office of Japan
amp: ampersands
NLP: natural language processing
PELT: pruned exact linear time
AWS: Amazon Web Service
DOV: degree of valence
LDA: Latent Dirichlet allocation

---

**Fig. 1.**
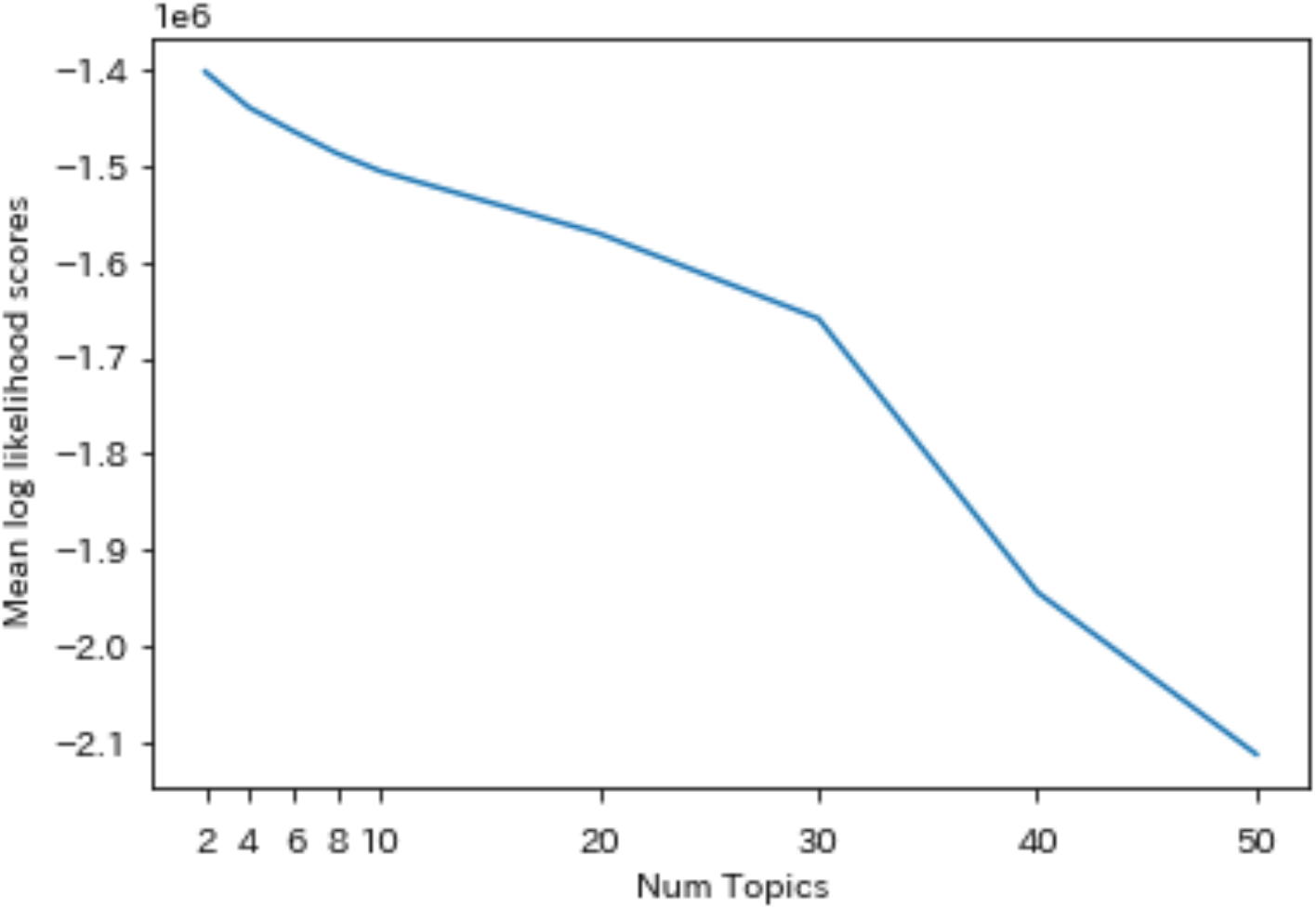
Mean Log likelihood scores for different LDA topic numbers using five-fold cross validation.

